# Safety and Efficacy of Daclatasvir with Sofosbuvir and Ribavirin in Hepatitis C Virus Infection: A Real World Experience from South Punjab, Pakistan

**DOI:** 10.1101/2021.10.23.21265410

**Authors:** Waseem Sarwar Malghani, Farooq Mohyud Din Chaudhary, Misbah Shahid, Ahsan Tameez-ud-din, Romaisa Malik, Asma Tameez Ud Din

## Abstract

**Background & Objectives:** Sofosbuvir (SOF) and daclatasvir (DCV) in combination with ribavirin (RBV) drastically changed the treatment scenario of chronic hepatitis C (CHC) patients, achieving remarkable efficacy and safety profile. Real world experience of SOF/DCV/RBV combination in this part of Asia was scant. This study aimed to evaluate the efficacy and safety of SOF/DCV/RBV combination to treat CHC patients at a tertiary care hospital in South Punjab.

**Methods:** Patients of CHC of any genotype were enrolled prospectively. They were treated with 12 weeks course of SOF/DCV/RBV combination. Effectiveness was evaluated by end of treatment response (ETR) and sustained virological response (SVR) at 12 and 24 weeks post-treatment. Adverse events were recorded for safety analysis.

**Results:** We analyzed data of 102 patients of CHC (40 males and 62 females). The mean age was 40.04 ± 10.22 years. Mean weight was 67.24 ± 11.78 kg, while mean body mass index (BMI) was 26.32 ± 4.58 kg/m^2^. Eighty patients belonged to low socio-economic status, while 22 belonged to middle socio-economic status. Sixty-four had a rural background, while 38 were from urban background. Seventy-four patients had no co-morbid condition; 16 (15.7%) had diabetes and 12 (11.8%) patients had co-morbid hypertension. Ninety percent of the patients did not have cirrhosis; 6% had compensated liver disease, while 4 % had decompensated liver disease. All the patients achieved undetectable HCV RNA at the end of treatment and 12 weeks after completion of treatment, while SVR at 24 weeks was achieved in 98% of patients. Only 2 patients discontinued treatment as a result of side effects. The most common side effects reported include fatigue, headache and fever.

**Conclusion:** CHC is a grave problem in developing countries like Pakistan. The SOF/DCV/RBV combination is very effective in eradicating CHC and has a very good side effect profile as well.

## Introduction

Hepatitis C affects a significant proportion of the world’s population with an estimated 71 million people having chronic hepatitis C infection (CHC) worldwide [1]. The burden of this disease in Pakistan is disturbingly high and it is second only to Egypt in terms of highest number of infections in a country. The reduction in this number is possible only by adopting a comprehensive test and treat strategy by the health authorities [2,3].

Since its discovery in 1975, there have been many treatment regimens against hepatitis C including interferon and ribavirin which were associated with low to intermediate cure rates [4]. The World Health Organization now recommends the use of direct acting antivirals (DAA) for 2-3 months which cure more than 90% of hepatitis C patients [4-6]. These drugs inhibit the viral proteins crucial for its replication hence quite literally stopping the virus in its tracks and paving the path towards patient recovery [7].

Daclatasvir (DCV) and Sofosbuvir (SOF) are two frequently recommended DAAs which have good efficacy and safety profile. The combination of these drugs is well tolerated and results in the achievement of a sustained viral response (SVR) in a significant proportion of the hepatitis C patients taking them [8]. Studies have shown that the addition of ribavirin (RBV) to this drug combination results in a sustained viral response even in the patients with liver cirrhosis [9]. The high efficacy and few side effects makes the combination of daclatasvir, sofosbuvir and ribavirin the treatment regimen of choice in this part of the world.

Owing to the alarmingly high prevalence of hepatitis C in Pakistan, there is a need of treating the patients with this new combination of drugs and publishing the data regarding its efficacy and safety. The objective of this study was to evaluate the efficacy and safety of SOF/DCV/RBV combination to treat CHC patients at a tertiary care hospital in South Punjab in order to improve the existing database regarding real world experience of this drug combination in this part of the world.

## Methods

This prospective study was conducted after getting approval from the Institutional Ethical Review Board (IRB No. 26880-935/NMU&H dated 10-10-2019), Nishtar Medical University Multan from October 2019 till August 2020. Informed consent was taken before incorporating patients into the study. The inclusion criteria included patients with chronic hepatitis C infection of any genotype and detectable HCV RNA on Polymerase Chain Reaction (PCR). Pregnant ladies, patients having any history of malignancy, patients younger than 18 years and patients with any co-infection of hepatitis B or human immunodeficiency virus (HIV) were excluded from the study. Patients were enrolled in the study and followed prospectively. At the start of treatment detailed history, examination, baseline investigations and an ultrasound abdomen were done of all the patients. They were treated with 12 weeks course of combination of Sofosbuvir 400mg, Daclatasvir 60mg and weight based ribavirin (1000mg in patients less than 75kg, while 1200mg in patients having eight more than 75kg).

During the 12 weeks course of SOF/DCV/RBV combination patients were regularly followed for the development of any side effects of the treatment. At the end of the course, effectiveness of treatment was evaluated by end of treatment response (ETR) and sustained virological response (SVR) at 12 and 24 weeks post-treatment by checking the HCV RNA levels by PCR. ETR was defines as undetectable HCV RNA at the end of treatment course. SVR at 12 and 24 weeks were defined as undetectable HCV RNA at 12 and 24 weeks after completion of treatment. Failure of treatment was defined as detectable HCV RNA at any of these strategic points.

## Results

We analyzed data of 102 patients of CHC. There were forty males and sixty-two females in our study population. The mean age was 40 years with a standard deviation of 10.22 years. Mean weight was 67.24 ± 11.78 kg, while mean body mass index (BMI) was 26.32 ± 4.58 kg/m^2^. Eighty patients belonged to low socio-economic status, while 22 belonged to middle socio-economic status. Sixty-four patients had a rural background, while 38 were from an urban background. Seventy-four patients had no co-morbid conditions; 16 (15.7%) had diabetes and 12 (11.8%) patients had co-morbid hypertension. The different demographic features of the study population are listed in Table 1.

**Table 1:** Demographic Characteristics of patients (n=102)

| Characteristic | N (%) |
| --- | --- |
| Age (years)<br>Mean ± Standard Deviation<br>Range | 40.04 ± 10.22<br>23 to 70 |
| Gender<br>Male<br>Female | 40 (39.2%)<br>62 (60.8%) |
| Weight (kg)<br>Mean ± Standard Deviation<br>Range | 67.24 ± 11.78<br>45 to 100 |
| Height (cm)<br>Mean ± Standard Deviation<br>Range | 156.72 ± 12.10<br>119 to 180 |
| Body Mass Index BMI (kg/m<sup>2</sup>)<br>Mean ± Standard Deviation<br>Range | 26.32 ± 4.58<br>15.5 to 36 |
| Follow up Time (months)<br>Mean ± Standard Deviation<br>Range | 11.20 ± 2.62<br>7 to 18 |
| Socioeconomic Status<br>Low<br>Middle | 80 (78.4%)<br>22 (21.6%) |
| Residential Status<br>Rural<br>Urban | 64 (62.7%)<br>38 (37.3%) |
| Co-morbid Conditions<br>No Co-morbid<br>Type 2 Diabetes Mellitus<br>Hypertension | 74 (72.5%)<br>16 (15.7%)<br>12 (11.8%) |

Table 2 explains the disease status of patients and treatment outcomes. Ninety percent of the patients did not have cirrhosis; 6% had compensated liver disease, while 4 % had decompensated liver disease. All the patients achieved undetectable HCV RNA at the end of treatment (ETR) and 12 weeks after completion of treatment (SVR at 12 weeks). However SVR at 24 weeks was achieved in 98% of patients. Only 2 patients discontinued treatment as a result of side effects. The most common side effect reported by the patients during the course of treatment was fatigue (43% of patients), followed by headache and fever. The different side effects according to their frequency are listed in Table 3. Twenty-seven percent of the patients reported no side effects during the treatment regimen.

**Table 2:** Disease Status and Treatment Outcome of Patients (n=102)

|  |  |
| --- | --- |
| Previous HCV Treatment Status |  |
| Treatment Naïve | 88 (86.3%) |
| Treatment Experienced (Interferon) | 14 (13.7%) |
| Disease Status |  |
| Non-Cirrhotic | 92 (90.2%) |
| Compensated Liver Disease (CTP-A) | 6 (5.9%) |
| Decompensated Liver Disease (CTP-B/C) | 4 (3.9%) |
| Treatment Outcome |  |
| ETR | 102 (100%) |
| SVR12 | 102 (100%) |
| SVR 24 | 100 (98%) |
| Treatment Discontinuation |  |
| No Discontinuation | 98 (96%) |
| Complete Treatment Discontinuation | 2 (2%) |
| Discontinued Ribavirin | 2 (2%) |

CTP: Child-Turcotte-Pugh; ETR: End of Treatment Response; SVR12: Sustained Virological Response at 12 weeks after completion of therapy; SVR24: Sustained Virological Response at 24 weeks after completion of therapy

**Table 3:** Side Effects reported by patients during the course of treatment.

| Side Effect | N (%) |
| --- | --- |
| Fatigue | 44 (43.1%) |
| Headache | 16 (15.7%) |
| Fever | 12 (11.8%) |
| Irritability | 8 (7.8%) |
| Insomnia | 4 (3.9%) |
| Pruritus | 2 (2%) |
| Hemolysis | 2 (2%) |
| Diarrhea | 2 (2%) |
| No side effects | 28 (27.5%) |

Some patients reported more than one side effect.

## Discussion

Chronic hepatitis C infection is a major cause of morbidity in the under developed parts of the world. Hepatitis C can be called the disease of poverty as the factors associated with its spread are largely due to poor hygiene, unemployment, low level of understanding and a general lack of education [10]. Historically, the impact of this disease in countries like Pakistan has been exacerbated to a debilitating extent due to the non-existence of a sufficiently effective and affordable treatment regimen. In the past few years, the introduction of the recently approved direct acting antivirals (DAA) have renewed a sense of optimism among the clinicians in Pakistan and it is hoped that by 2030, a significant reduction in the hepatitis C prevalence will be achieved [2,3].

This study was designed to improve the understanding about the safety and efficacy of SOF/DCV/RBV combination in CHC patients. The number of patients included in our study was 102 with the mean age of 40 years. Almost 60 percent of the study population consisted of females. Our study was demographically comparable to other studies from Pakistan [11,12]. Most of the patients (78.4%) had a low socioeconomic status. Low level of education and lack of basic hygienic measures is a major cause of spread of this disease in our country. Reuse of injection needles and improper disposal of shaving blades are considered two of the biggest factors behind the rapid dissemination of hepatitis C infection [13]. Carefully designed public health campaigns and strict legislation are needed to tackle this serious issue in an effective way.

More than 86% of the patients were treatment naïve while 13.7% had a past treatment experience with Interferon. All the patients achieved undetectable HCV RNA at the end of treatment (ETR). This was similar to the results reported by Jamil et al where 98.7% of patients achieved ETR [12]. Comparable results were reported by researchers from other countries [14]. In our study, SVR at 12 weeks was achieved in all 102 patients while SVR at 24 weeks was reported for 98% of the patients. Lionetti et al reported almost identical results with a 100% of the treatment group receiving SOF/DCV/RBV achieving SVR at 12 weeks and 90.4% of the patients receiving SOF/DCV only having undetectable HCV RNA after 12 weeks [14]. A study conducted in Egypt reported 96% SVR at 12 weeks in patients using a combination of SOF/DCV [15]. The data regarding the efficacy of these drugs has been encouraging and studies from around the globe have replicated such positive results [15,16].

The side effects of this drug combination reported by the participants were mild and only 2 patients discontinued the complete treatment regimen. The most common side effect was fatigue (43.1%) followed by headache (15.7%) and fever (11.8%). Other less common side effects included irritability, insomnia, pruritus, hemolysis and diarrhea. Headache and fatigue were reported to be the most common side effects associated with the SOF/DCV drug combination in a study by Nelson et al [17]. In contrast, Lionetti et al reported anemia as the most common adverse event in patients receiving RBV along with the other two drugs leading to a dose reduction of RBV in some patients [14].

There are quite a few studies that corroborate the findings of this article concerning the high efficacy and favorable safety profile of the direct acting antivirals (used with or without ribavirin) [15-17]. The adverse events leading to discontinuation or death are very rare. This makes the combination of these drugs the drug of choice for combating hepatitis C disease in Pakistan and the ultimate weapon against this highly prevalent and dangerous disease.

There were some limitations to our study. Genotype testing could not be performed due to the issues regarding the availability of specific laboratory equipment and affordability of the patients. Side effects on different lab parameters at the end of treatment were not measured in many patients due to loss to follow up.

## Conclusion

Chronic Hepatitic C is a grave problem in developing countries like Pakistan. The introduction of SOF/DCV/RBV combination in government hospitals have drastically reduced the cost of treatment of this illness. The high efficacy, tolerable side effects and low cost of treatment give hope that someday we can get rid of this menace from Pakistan.

## Data Availability

All data produced in the present work are contained in the manuscript.

